# Systematic review of international guidelines for tracheostomy in COVID-19 patients

**DOI:** 10.1101/2020.04.26.20080242

**Authors:** Carlos M. Chiesa-Estomba, Jérome R. Lechien, Christian Calvo-Henríquez, Nicolas Fakhry, Petros D. Karkos, Shazia Peer, Jon A. Sistiaga-Suarez, José A. Gónzalez-García, Giovanni Cammaroto, Miguel Mayo-Yánez, Pablo Parente-Arias, Sven Saussez, Tareck Ayad

## Abstract

At this moment, the world leaves under the SARS-CoV-2 outbreak pandemic. As Otolaryngologists - Head & Neck Surgeons, we need to perform and participate in examinations and procedures within the head and neck region and airway that are at particularly high risk of exposure and infection because of aerosol and droplet contamination. One of those surgical procedures on demand at this moment is tracheostomy, due the increasing admission in ICU departments and the increased need of ventilatory support secondary to respiratory distress syndrome. This review of international guidelines for tracheostomy in COVID-19 infected patients, aiming to summarize in a systematic way the available recommendations from 18 guidelines from all over the world.

## Introduction

Since the first described cases of severe acute respiratory syndrome coronavirus- 2 (SARS-CoV-2), identified in the Hubei region of China in December 2019, Coronavirus is spreading rapidly across countries.^1^

On March11 2020, the World Health Organization (WHO) finally declared the SARS-CoV-2 as an outbreak pandemic due to the increasing number of cases around the world.^2^ Patients with SARS-CoV-2 infection can develop the also known as coronavirus disease 2019 (COVID-19), which has resulted in high rates of emergency visit, hospitalization and intensive care unit (ICU) admissions.^3^ Up to April the 10th 2020, a total of 1.521.252 patients had tested positive for the new SARS-CoV-2 coronavirus worldwide and 92.798 (6.1%) died.^4^ However, reported mortality could vary because of differences in proportions of elderly in the studied populations, differences in the prevalence of comorbidities in different areas or countries and differences in country resources and criteria for testing of the population.^5^

Despite Information on the incidence of the disease, clinical characteristics of the critically ill patients diagnosed with COVID-19 are still limited. According to the Johns Hopkins Dashboard the infection affects predominantly in people of 30-79 years-old (87%). Almost 81% are asymptomatic or experience mild symptoms, whereas 15% of patients suffer severe symptoms requiring hospitalization and between 3-15 % will need respiratory support in an intensive care unit (ICU) setting with mechanical ventilation or extracorporeal membrane oxygenation (ECMO).^6-8^

As Otolaryngologists - Head & Neck Surgeons, we need to perform and participate in examinations and procedures within the head and neck region and airway that are at particularly high risk of exposure and infection because of aerosol and droplet contamination.^9^ One of those surgical procedures on demand at this moment is tracheostomy, due the increasing admission in ICU departments and the increased need of ventilatory support secondary to respiratory distress syndrome. In this respect, it is of paramount importance to establish tracheostomy guidelines that focus equally on both patient’s and health care team’s well-being during the COVID-19 pandemic in order to minimize risk of viral exposure.

This review of international guidelines for tracheostomy in COVID-19 infected patients is an initiative of the “*Young Otolaryngologist Group of the International Federation of Otolaryngologic Societies”* (YO-IFOS), aiming to summarize in a systematic way the available recommendations.

## Methods

The systematic review was performed in accordance with PRISMA guidelines.^10^ This review involved a systematic search of the electronic databases MEDLINE/PUBMED, Google Scholar, the Chinese SinoMed (http://www.sinomed.ac.cn/zh/), Ovid Medline, Scielo, Embase, Scopus, and the Database of Abstracts of Reviews of Effects and directly contacting different ENT-HNS societies and institutions across the world. Guidelines and recommendations from January 2020 to April 2020 were included. No language restriction was applied to the search strategy. Search terms (medical subject headings or keywords) included: “tracheotomy”, “tracheostomy,” “SARS CoV-2”, “COVID-19”, “guidelines/practice guidelines/clinical guidelines,” “consensus,” “ENT,” and “Head & Neck surgery.” Titles and abstracts were screened by two investigators (C.M.C.E and T.A) to discard irrelevant publications.

Inclusion criteria for the systematic review were based on the population, intervention, comparison, and outcomes according to (PICO) framework.^11^

### Population

Inclusion criteria consisted of clinical guidelines comprehensively reporting Severe COVID-19 patients treatment strategies.

### Intervention and comparison

Intervention and comparison were defined according to the need for tracheostomy, COVID-19 status, indications for tracheostomy, measures of protection, type of tracheostomy technique, diathermy or not, surgical setting, post-tracheostomy care.

### Outcome

The primary outcome was to describe a set of recommendations to improve tracheostomy safety.

Common questions related to surgical safety, indications, COVID-19 status, patients’ comorbidities, standard personal protective equipment (PPE), setting, methods of dissection, type of tracheostomy and post-tracheostomy care were reviewed. All searches were completed in April 13, 2020.

Guidelines Credibility, as measured by whether the guidelines were developed, cited by subsequent guidelines or by other publications regarding tracheostomy in COVID-19 patients was assessed. The most recent version or update of each clinical guideline was included and analyzed.

To assess the methodological quality of guidelines included The AGREE II (Appraisal of Guidelines for Research and Evaluation II) instrument was used.^12^ The quality assessment of all included clinical practice guidelines was performed by four appraisers independently (C.C.E, J.A.S, J.A.G.G & T.A). We defined a cut□off of 50%, and scores above this were considered acceptable. To measure interobserver agreement across the ordinal categories of the AGREE-II ratings, a weighted kappa was calculated using SPSS V.21.0. The degrees of agreement were graded as minor (≤0.20), fair (0.21– 0.40), moderate (0.41–0.60), substantial (0.61–0.80) and almost perfect (≥0.81).

## Results

The electronic search strategy identified 18 guidelines for analysis, 17 about tracheostomy including 1 guideline from Oceania (Australian and New Zealand),^13^ 6 from North America (1 from Canada and 5 from USA),^14-219^ 3 from the United Kingdom,^20-22^ 4 from Europe (2 Spanish, 1 French and 1 Italian),^23-26^ 2 from Africa (Both from South Africa)^27,28^ and 1 from Asia (1 Singapore),^29^ all of them cited in Table 1. Additionally, two guidelines focusing specifically on post-tracheostomy care were included. (1 from France and 1 from Spain) (Table 2).^25,30^ Data from other Asian countries, Middle East, South, Central America and the Caribbean were not available online. These guidelines were examined and compared, results are summarized in table 1 and 2 and figure 1.

**Table 1.**
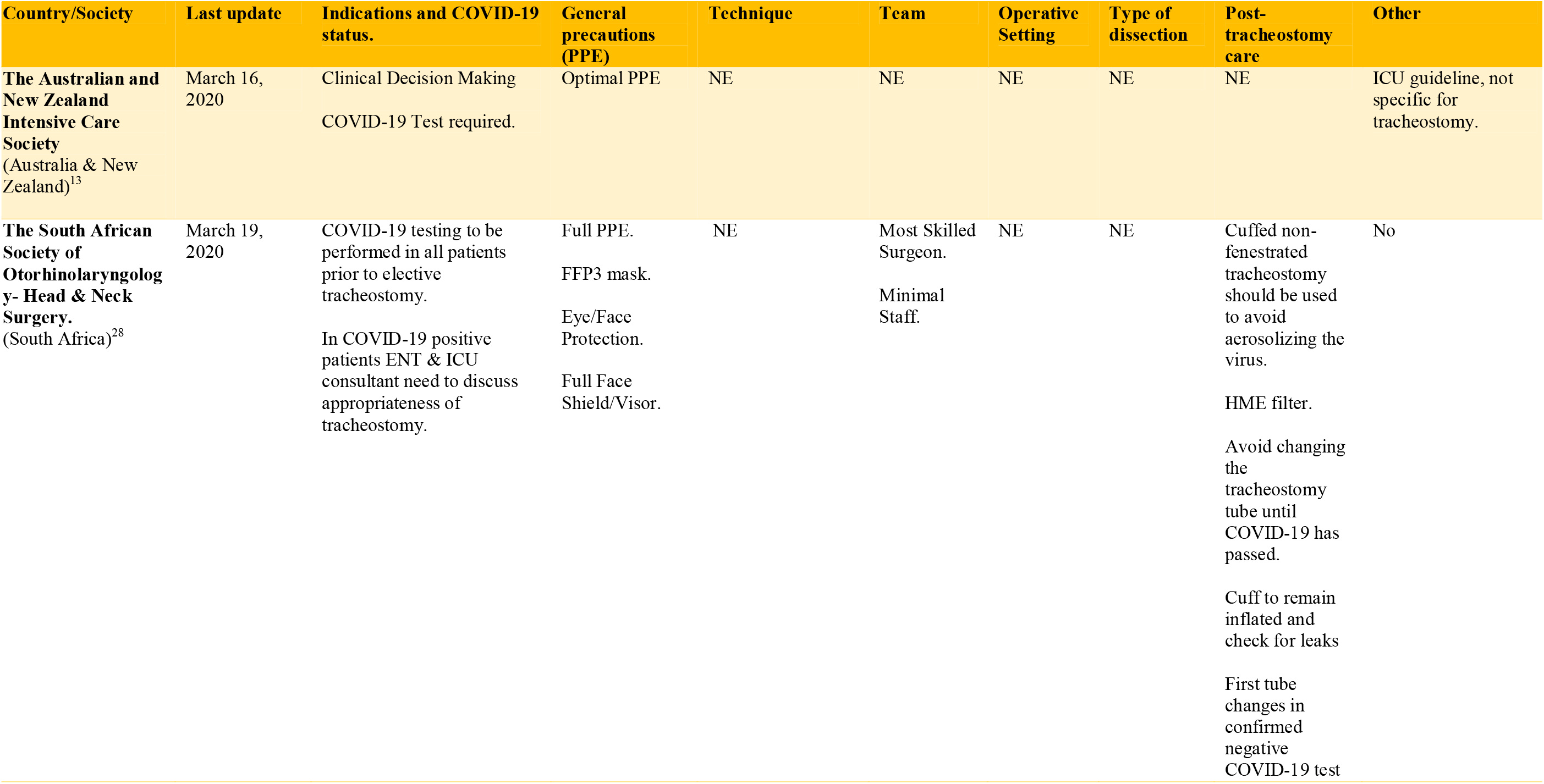
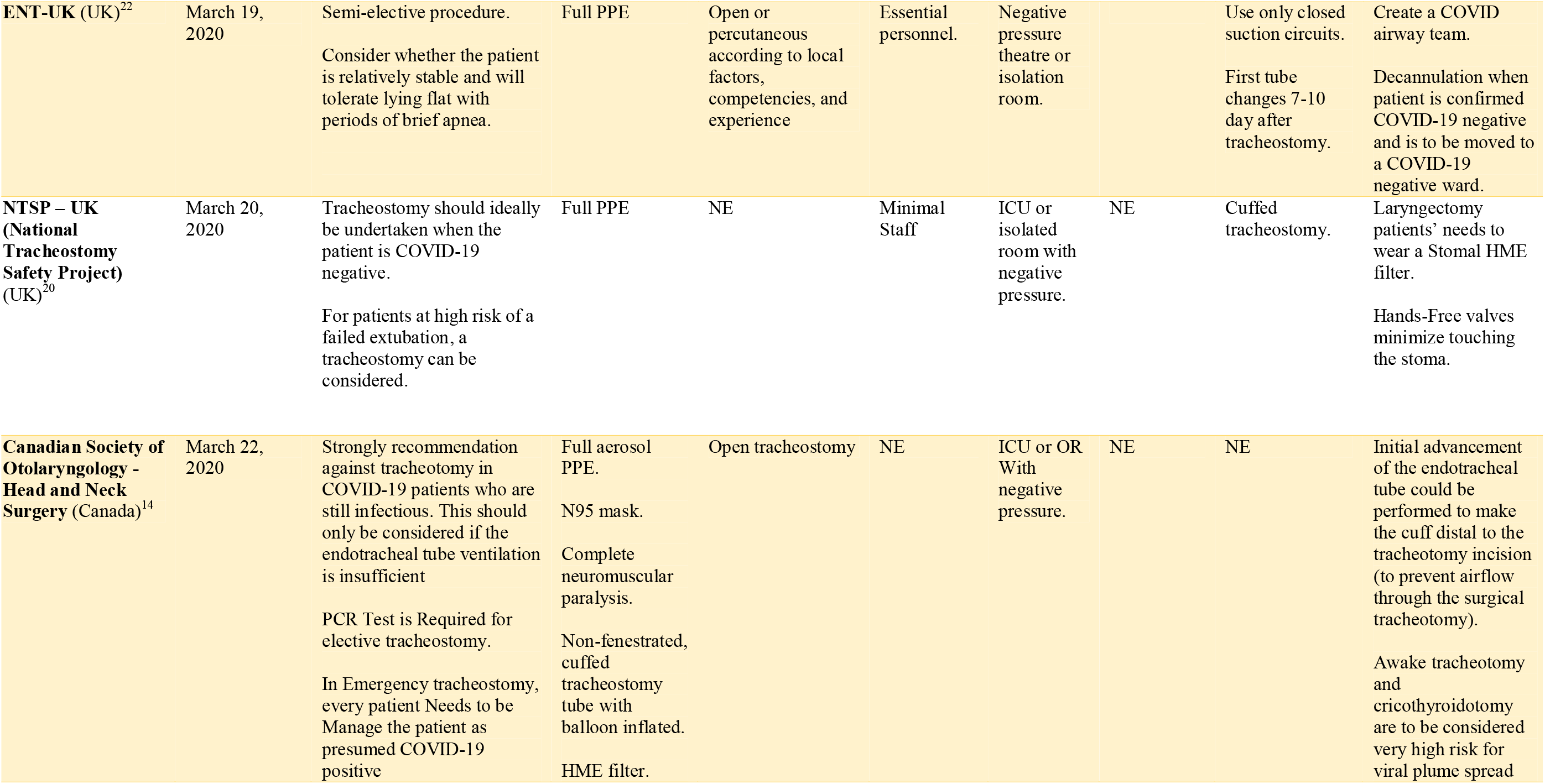
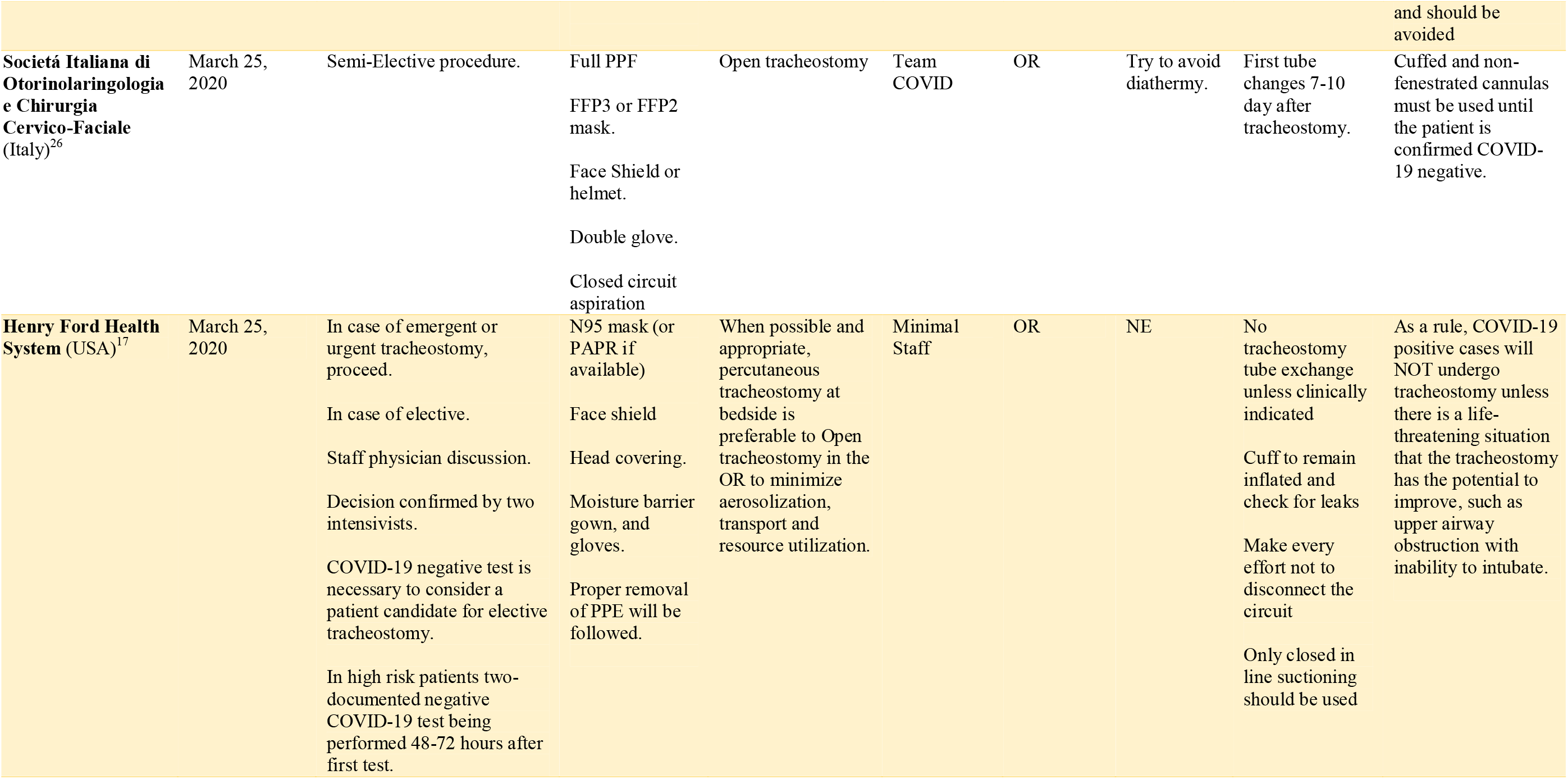
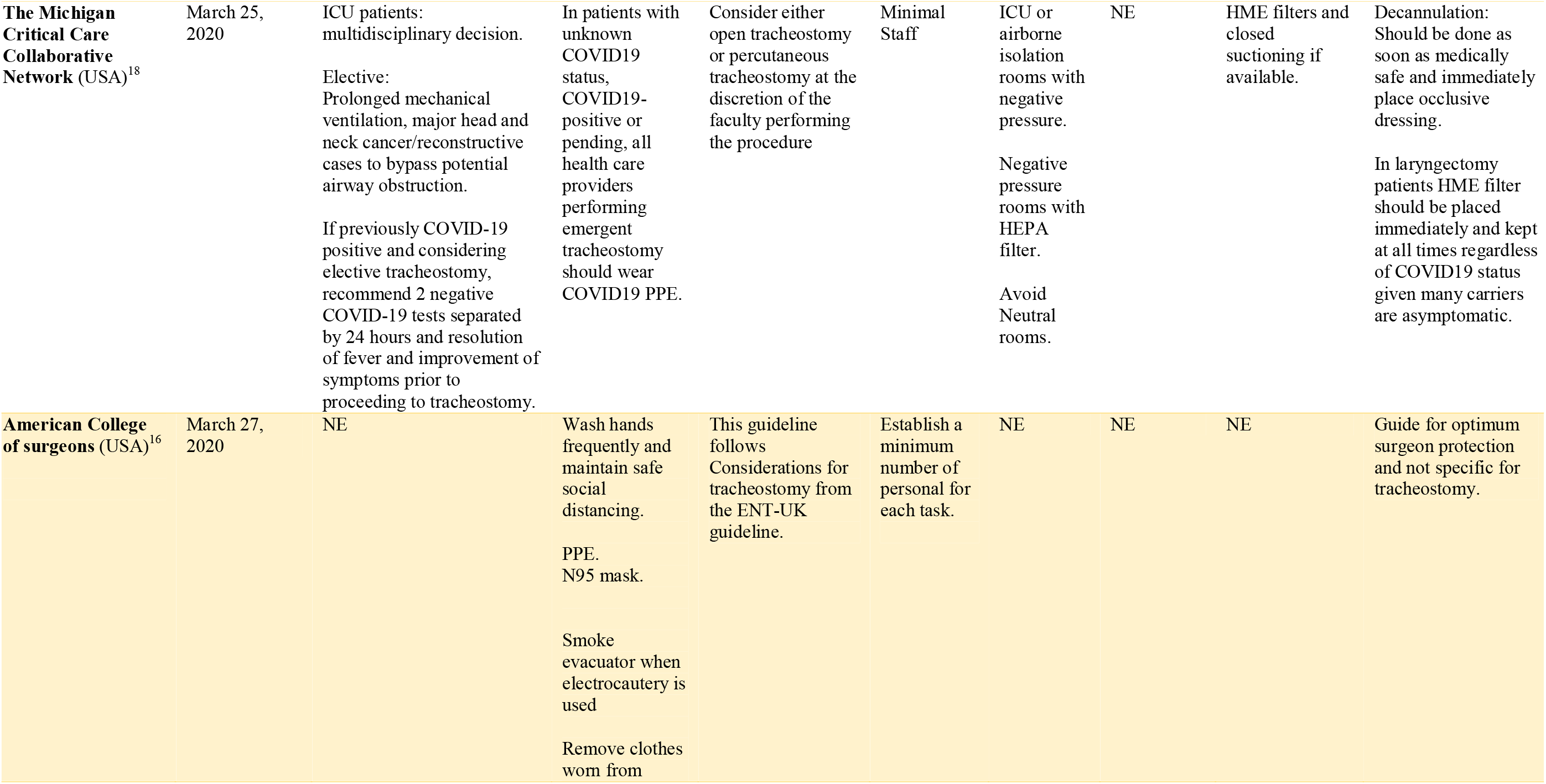
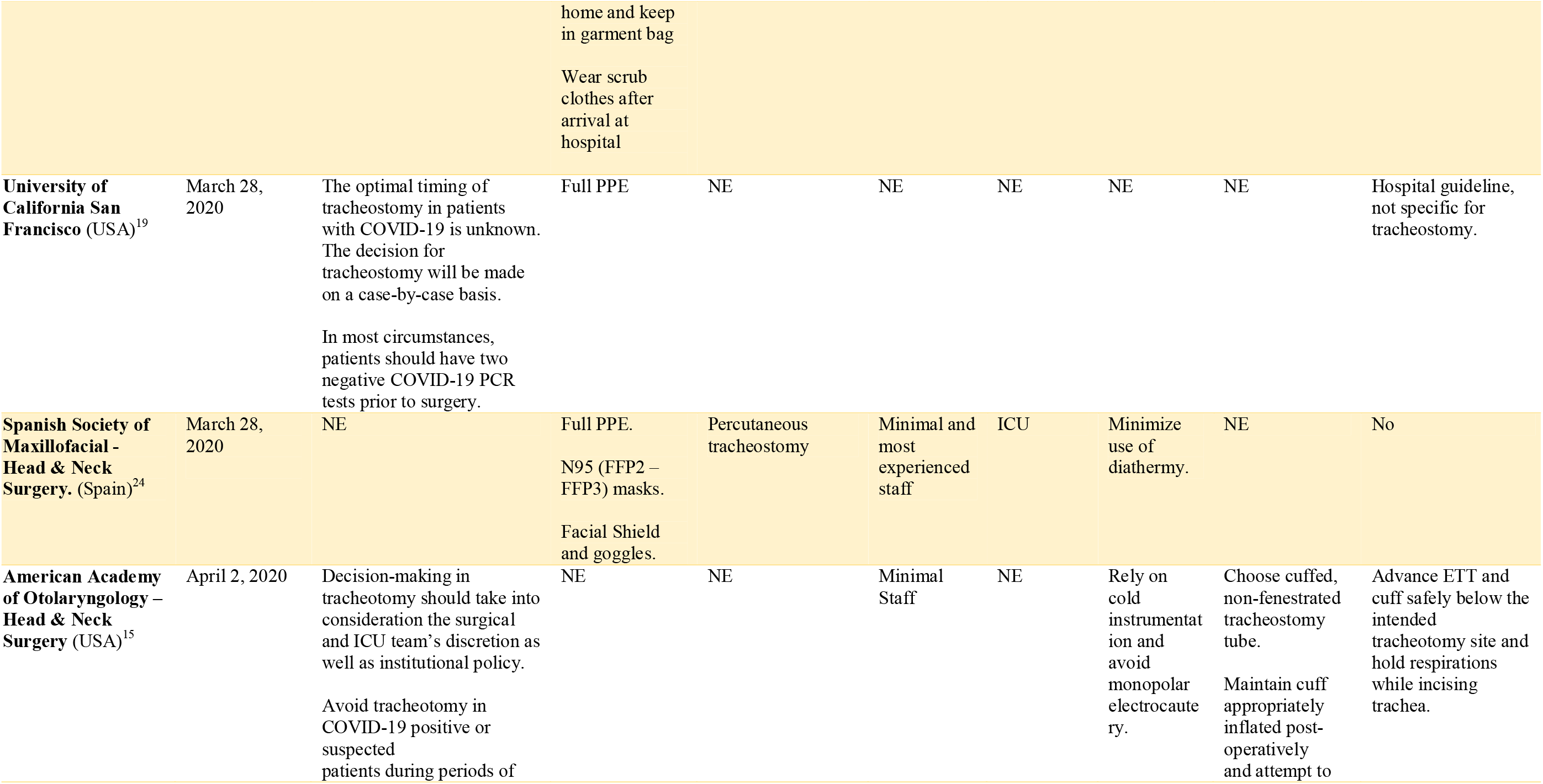
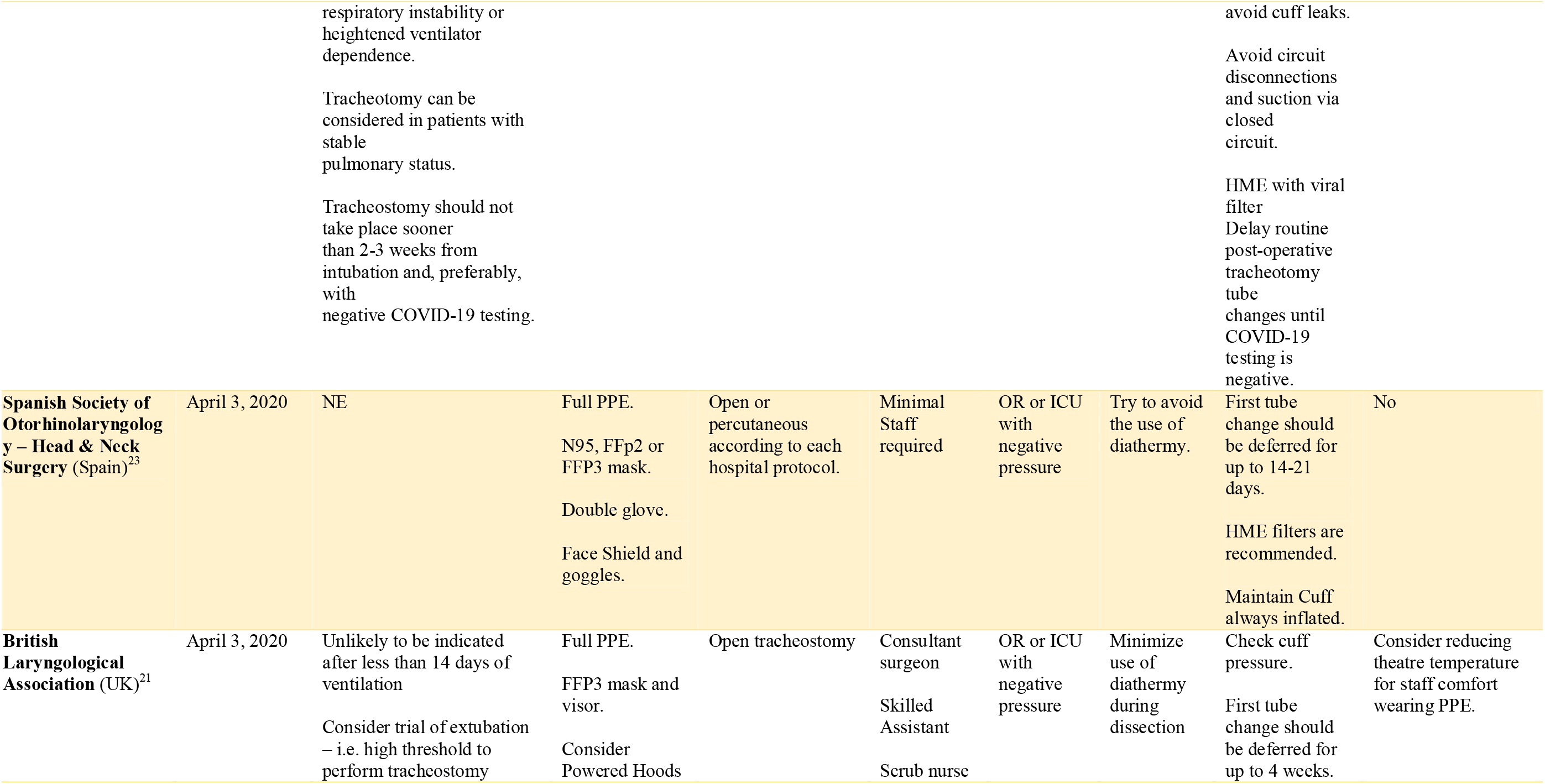
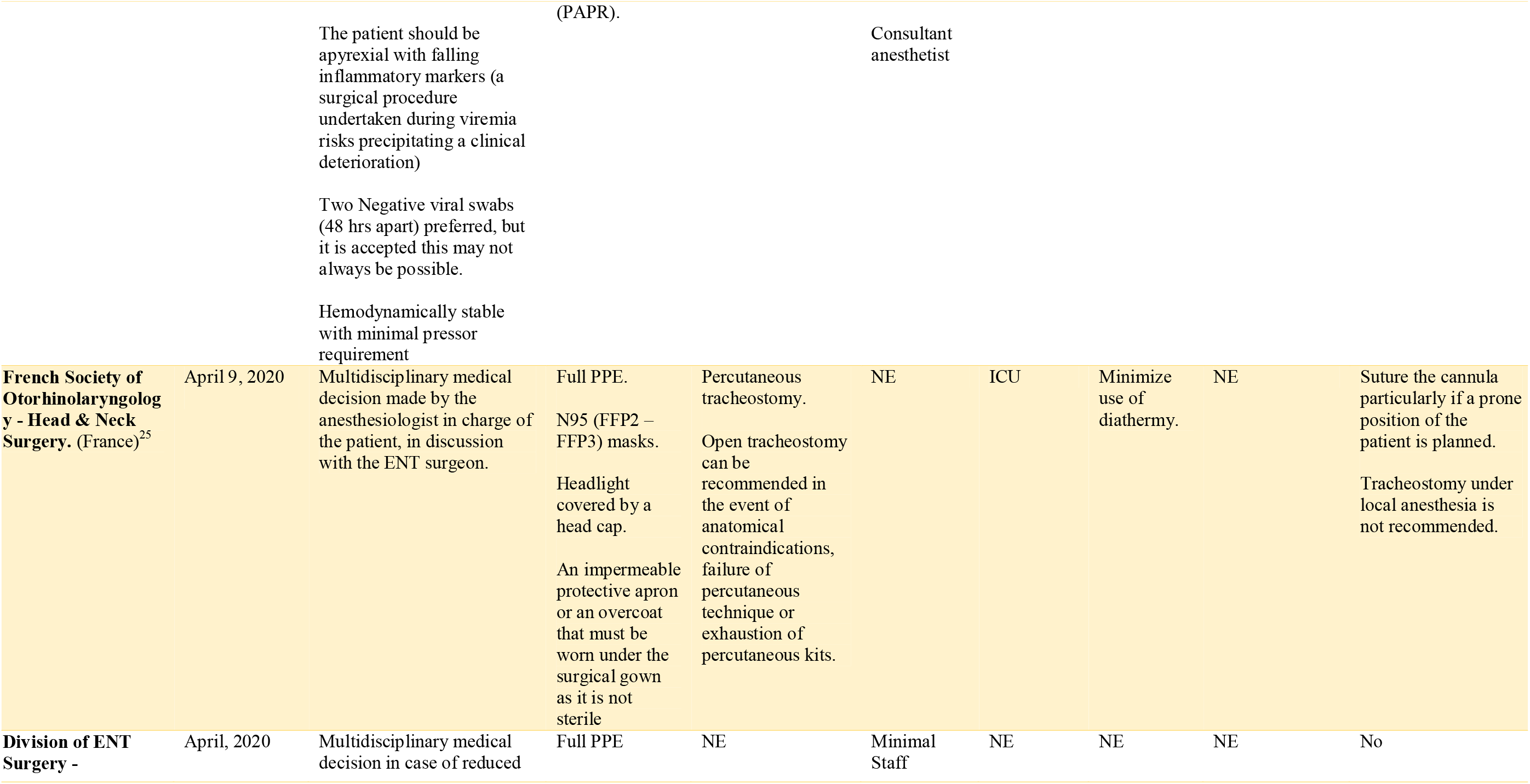
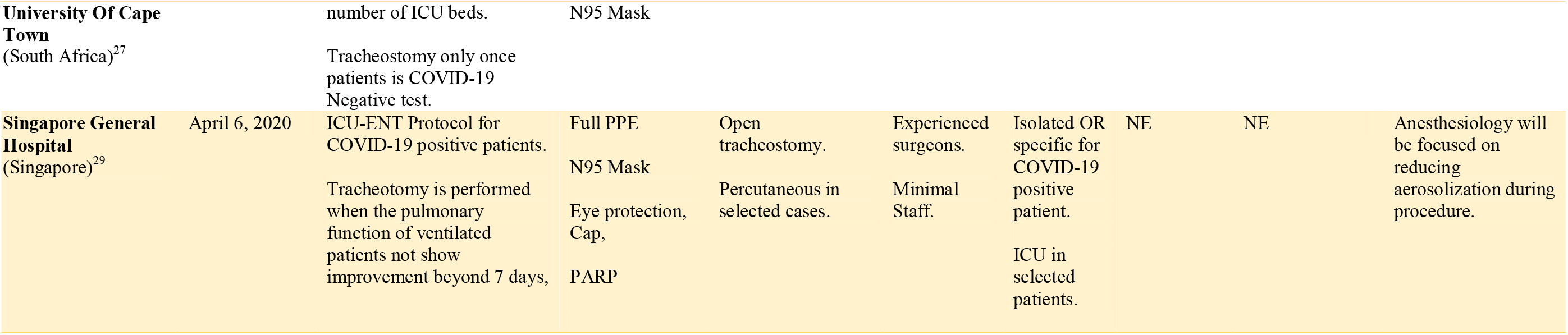
Worldwide guideline comparison. OR (Operating Room), COVID-19 (Coronavirus disease 2019), ICU (Intensive Care Unit), PPE (Personal Protective Equipment), NE (Not especified), PARP (Powered air-purifying respirator)

**Table 2.** Tracheostomy care guidelines. HME (Humidity-Moisture Exchange), HEPA ((High Efficiency Particulate Air), NE (Not specified).

| Country | Last Update | COVID-19 Status | Airway protection for patients | Tracheal tube changes by healthcare workers | Voice Prosthesis changes | Other |
| --- | --- | --- | --- | --- | --- | --- |
| **Spanish Society of Otorhinolaryngology – Head & Neck Surgery.**<br>(Spain)<sup>30</sup> | March 26, 2020 | COVID-19 Negative | HME filter, surgical mask over the tracheal stoma when is indicated or HEPA filter. | Basic PPE (Gloves, Goggles, Surgical mask or FFP2, gown) | Basic PPE | No |
|  |  | COVID-19 Suspected or Positive. | HME filter or HEPA filter. | Full PPE. | Full PPE | Aspiration on close suction system. |
| **French Society of Otorhinolaryngology - Head & Neck Surgery.**<br>(France)<sup>25</sup> | April 9, 2020 | Any patient should be considered as potentially infected | NE | Apron or surgical gown.<br>Forehead protection to protect skin exposure.<br>Gloves.<br>Goggles.<br>FFP2 – N95 Mask. | NE | If the patient is ventilated on the tracheostomy cannula, ask the anesthesiologist to sedate the patient and perform a neuromuscular block to reduce any risk of coughing during the change of the cannula. |

### Appraisal of guidelines

The AGREE-II domain percentages for each guideline are shown in table 3. None of those selected guidelines stated the methods used in the literature search, the quality of the evidence, and the strengths of the recommendations reported.

**Table 3.** Guideline assessment according to the AGREE-II Instrument

| Guideline organization or society | Scope and Purpose | Stakeholder Involvement | Rigor of development | Clarity and Presentation | Applicability | Editorial Independence | Mean domain scores (%) | Agreement between appraisers (Kappa 95% CI) |
| --- | --- | --- | --- | --- | --- | --- | --- | --- |
| **The Australian and New Zealand Intensive Care Society**<br>(Australia & New Zealand)<sup>13</sup> | 66.6 | 63.8 | 57 | 61.1 | 69.4 | 75 | 65,4 | 0.63 (0.59 to 0.68) |
| **The South African Society of Otorhinolaryngology-Head & Neck Surgery.**<br>(South Africa)<sup>28</sup> | 94,6 | 72.2 | 55 | 83.3 | 80.5 | 83 | 78,1 | 0.77 (0.63 to 0.87) |
| **ENT-UK (UK)**<sup>22</sup> | 100 | 70.5 | 59,5 | 88.8 | 91.6 | 83 | 82,2 | 0.92 (0.72 to 0.97) |
| **NTSP – UK (National Tracheostomy Safety Project) (UK)**<sup>20</sup> | 94,6 | 76.5 | 55 | 83.3 | 86.1 | 70.8 | 77,7 | 0.84 (0.77 to 0.90) |
| **Canadian Society of Otolaryngology - Head and Neck Surgery** (Canada)<sup>14</sup> | 100 | 65 | 57 | 86.1 | 88.8 | 87.5 | 77 | 0.81 (0.69 to 0.89) |
| **Società Italiana di Otorinolaringologia e Chirurgia Cervico-Faciale** (Italy)<sup>26</sup> | 100 | 62.2 | 52,3 | 83.3 | 82.1 | 75 | 75,8 | 0.73 (0.54 to 0.97) |
| **Henry Ford Health System** (USA)<sup>17</sup> | 94,6 | 72.2 | 52,4 | 83.3 | 86.1 | 70.8 | 76 | 0.77 (0.58 to 0.91) |
| **The Michigan Critical Care Collaborative Network** (USA)<sup>18</sup> | 94,6 | 66.6 | 55 | 80.5 | 83.3 | 70.8 | 75,1 | 0.75 (0.54 to 0.83) |
| **American College of surgeons (USA)**<sup>16</sup> | 66.6 | 69,4 | 50 | 61,1 | 77 | 75 | 66.5 | 0.84 (0.67 to 0.93) |
| **University of California San Francisco (USA)**<sup>19</sup> | 94,6 | 72.2 | 51.4 | 77 | 83.3 | 70.8 | 74.8 | 0.92 (0.73 to 0.99) |
| **Spanish Society of Maxillofacial - Head & Neck Surgery. (Spain)**<sup>24</sup> | 94,6 | 72.2 | 52.4 | 83.3 | 86.1 | 83 | 78.6 | 0.47 (0.39 to 0.56) |
| **American Academy of Otolaryngology – Head & Neck Surgery (USA)**<sup>15</sup> | 100 | 74.5 | 57 | 86.1 | 88.8 | 83 | 81.5 | 0.86 (0.71 to 0.96) |
| **Spanish Society of Otorhinolaryngology – Head & Neck Surgery (Spain)**<sup>23</sup> | 100 | 72.2 | 52.4 | 86.1 | 88.8 | 81 | 80 | 0.71 (0.65 to 0.84) |
| **British Laryngological Association (UK)**<sup>21</sup> | 100 | 78.5 | 59.5 | 88.8 | 91.6 | 83 | 83.5 | 0.81 (0.71 to 0.89) |
| **French Society of Otorhinolaryngology - Head & Neck Surgery. (France)**<sup>25</sup> | 100 | 72.5 | 52.4 | 83.3 | 86.1 | 87.5 | 80.3 | 0.79 (0.71 to 0.93) |
| **Division of ENT Surgery - University Of Cape Town (South Africa)**<sup>27</sup> | 94,6 | 72.2 | 55 | 83.3 | 83.3 | 87.5 | 79,3.6 | 0.78 (0.69 to 0.84) |
| **Singapore General Hospital (Singapore)**<sup>29</sup> | 100 | 72.2 | 52,4 | 86.1 | 86.1 | 87.5 | 80,7 | 0.87 (0.72 to 0.96) |

#### Scope and Purpose

With this quality domain, the aim of the guideline under scrutiny is determined. A total of 8 guidelines obtained a score of 100%,^14,15,21-26,29^ 7 a score >80%,^17-20,24,27,28^ and 2 a score <70^13,16^. Therefore, 15 of the reviewed guidelines had a clearly described scope and purpose.

#### Stakeholder Involvement

No guidelines scored > 80%,^13,30^ with that no guidelines including the views and preferences of the patients and users. Most guidelines do not provide details about the participants during the developmental process. In general, the target audience is generally specified in the guideline procedure.

#### Rigor of Development

Details about the research strategy, evidence rating scales and evidence selection criteria were not described in any guidelines.^13-29^ The update was indicated in 2 guidelines.^15,23^

#### Clarity of Presentation

All guidelines provided a clear presentation and summary of recommendations. One of them offered an algorithm.^26^

#### Applicability

All guidelines offered elements regarding applicability in the context of operative room or ICU setting, and protective measures were provided to healthcare workers. Also, elements concerning implementation were described on each guideline. Just one guideline provides recommendations in a resource-constrained setting.^27^

#### Editorial Independence

The overall mean domain score and SD were 79 ± 6.3%. No guidelines provided information about interest or financing. However, almost all come from institutions or national societies and this information is not relevant for tracheostomy in the context of the pandemic.

#### Overall Assessment

Fifteen (88%) guidelines were assessed and recommended for use during COVID-19 pandemic, since their quality scores 6 or more,^14,15,17-29^ representing good-quality guidelines. The 2 remaining guidelines had scores between 5 and 6.^13,16^ These guidelines were recommended for COVID-19 attention, but not as tracheostomy guidelines.

The mean scores (range; SD) for the domains were: scope and purpose 93.8% (66.6%–100%; SD 10.22); stakeholder involvement 70.8% (62.2%–78.5%; SD 4.18); rigor of development 54.4% (51,4%–59,5%; SD 2.61); clarity of presentation 81.4% (61,1%–88,8%; 7.91); applicability 84.6% (69,4%–91,6%; SD 5.30) and editorial independence 79.6% (75%–87.5%; 6.33). The kappa values ranged from 0.47 (95% CI 0.39 to 0.56) to very good 0.92 (95% CI 0.73 to 0.99). The overall inter-rater agreement was intraclass correlation=0.89 (95% CI 0.81 to 0.94), indicating very good strength of agreement.

### Statements

The following statements reflects the current practice of surgical airway management in patient suffering a severe COVID-19 infection. Given the dynamic nature of scientific information and technology and the emergence of new data during and after the outbreak, the information herein presented should be managed carefully. Periodic reviews, updates, and revisions are required and expected. Tracheostomy surgical steps and technical notes are beyond the scope of this paper.

#### 1. Is it relevant to know COVID-19 status before performing a tracheostomy?

The availability to test patients differs among countries, even among each region within every country. Ideally, being able to assess COVID-19 status prior to performing a tracheostomy, is of paramount importance. PCR or serology tests are scarce due to the outbreak and the limited resources in some settings needs to be taken into consideration. Moreover, data regarding viral load in the upper airway (nasopharynx and oropharynx) is confusing. Early data suggest a higher viral load in the early phase of the disease, between days 9 to15.^31,32^ However, new evidence suggests viral loads detection in secretions up to 2-3 weeks, after onset of symptoms.^33^

According to guidelines reviewed; some recommend performing a test before any elective tracheostomy.^13-21,27,28^ Furthermore, for SARS-CoV-2 positive patients, there is a strong recommendation against performing a tracheostomy in patients who are infectious and have a high viral burden. One guidelines specifies that during the pandemic all patients should be considered positive (until proven otherwise) and full PPE needs to be used during each procedure.^18^ Other guidelines do not specify or mention the need of a recent test prior to a tracheostomy.^23-26^ All guidelines recommend, as expected, proceeding in case of emergency or crash tracheostomy regardless of COVID-19 status, using all full PPE.^13-29^

#### 2. What precautions and measures are needed to perform a tracheostomy?

During the outbreak, barrier precautions are critical in order to protect healthcare workers from contamination or infection. All the consulted guidelines recommends a full PPE due to the high risk of infection, including N95 or N99 (USA), FFP2 or FFP3 (Europe) mask, the use of double gloves, goggles or eye protection, Powered air-purifying respirator (PARP), face shield and an apron or gown.^13-29^ Careful hand hygiene is recommended with hydroalcoholic solution. After every tracheostomy, careful PPE removal following the sequential protocol is important to avoid contamination. Also, a designated person to closely supervised this step is mandatory.^34^

#### 3. What is the role of patients’ comorbidities?

It is relevant to understand that COVID-19 patients with some comorbidities have a poor prognosis.^35^ Identifying the most important risk groups is essential during the decision-making process. A recent meta-analysis identified hypertension, diabetes, chronic obstructive pulmonary disease, cardiovascular disease, and cerebrovascular disease as significant risk factors for COVID-19 patients.^36^ Also, the Italian experience have informed us that elderly patients had a higher mortality compared to younger patients (36%vs 15%; difference, 21% [95%CI, 17%-26%]; P < .001).^37^

#### 4. What are the risks related to performing a tracheostomy in COVID-19 positive patients?

Open and percutaneous tracheostomy represent an aerosol generating procedures (AGP), related to an increases in potential viral exposure for the healthcare team involved.^38^ It is attributed to aerosolized viral particles, which can stay for up to 3 hours.^39^ Furthermore, after tracheostomy, there is an increased potential for virus exposure to the ICU team who perform several bedside procedures from simple examination to suctioning, dressing changes and other routine post-tracheotomy care.^15^

According to Wuhan epidemic data, affected health workers represented 3.8% of the infected patients and 14.8% of them suffered a severe disease, with an overall mortality rate of 0.6%.^6,40,41^ Also, In Italy and Spain, 15-20% of responding healthcare workers were infected, and some have died.^42^

#### 5. Is there any evidence regarding indications and timing for elective tracheostomy?

Indications and timing represent the most controversial issue. Is important to highlight that the indication comes from the ICU <u>clinicians</u> or anesthesiologists in charge of the patients and there are lot of parameters that need to be taken into consideration. It’s necessary to establish a multidisciplinary team (ICU doctor, Anesthesiologist, ENT or Head & Neck Surgeon) to discuss every single case. Our findings present a high variability in national guidelines and protocols guidelines. European guidelines propose a more aggressive approach (early tracheostomy)^23,24,25^, maybe attributed the high volume of cases at this moment of the outbreak. By contrast British, North American, Singaporean and South-African guidelines propose a more conservative approach, suggesting waiting for at least 14 days of ventilation or a COVID-19 negative test (PCR or Two negative pharyngeal swabs) before performing an elective tracheostomy. They also recommend avoiding tracheostomy in COVID-19 positive or suspected patients during periods of respiratory instability or heightened ventilator dependence.^13-22,27-29^

Looking at previous reports, benefits of performing an early tracheotomy in critically ill COVID-19 patients are unclear and based on the SARS-CoV-1 outbreak with a similar coronavirus, demonstrated that the need for mechanical ventilation was associated with a 46% mortality.^43^ Also, during the SARS-CoV-1 outbreak doctors were unable to identify the exact time-point when afflicted patients either improve, remain stable, or progress toward death due to pulmonary complications. As the mean time from onset to death was 23.7 days, there was a very low potential benefit of tracheotomy prior to this time.^44^

Early evidence from Wuhan shows that the time from hospital admission to death was log□normally distributed with mean 11□days and median 5□days. However, this data not reach statically significance (*p* = 0.56).^45^ Givi *et al*. suggests delaying most tracheostomy procedures beyond 14 days, first of all given the high infectious risks during the procedure, arguing that early phases of COVID-19 infection may be related to a higher viral load. Secondly, given that after 14 days the likelihood of recovery is higher, and the ventilator weaning has become the primary goal of care.^9^ A recent systematic review suggested that early tracheostomy, performed in the first 7 days after orotracheal intubation, is associated with a reduction of mechanical ventilation duration, mortality rate and length of stay in ICU irrespective of reason for admission.^46^ By contrast, a previous randomized clinical trial, did not show an improved mortality rate or reduced length of ICU stay in patients on mechanical ventilation that underwent early tracheostomy.^47^ Furthermore, a meta-analysis of randomized controlled clinical trials with trial sequential analysis performed by Ruohui *et al*, compared with late tracheotomy, early tracheotomy presented a lower incidence of ventilator-associated pneumonia, shorter duration of mechanical ventilation, and shorter ICU stay. However, trial sequential analysis (TSA), a kind of cumulative meta-analysis, indicated that the evidence was unreliable and inconclusive.^48^

However, more data from COVID-19 patients is needed to compare both early versus late and suggest proper timing to perform a tracheostomy. Meanwhile seems prudent to evaluate individually prognosis factors before doing a procedure that could expose healthcare workers.

#### 6. Percutaneous tracheostomy versus surgical tracheostomy, what is the best option to choose?

After reviewing the guidelines, we found recommendations against and in favor of both techniques. Six guidelines did not specify the type of technique recommended,^13,15,19,20,28,29^ 3 guidelines recommend open tracheostomy,^14,21,26^ 1 recommends percutaneous tracheostomy^17^ and 6 recommend both techniques.^16,18,22,23,25,29^ Nevertheless, data about aerosol diffusion or droplets during percutaneous or open tracheostomy are not available across the literature. Low-level evidence from SARS-CoV1 outbreak was in favor of open tracheostomies (OT).^49,50^

Percutaneous tracheostomy (PT) hypothetically involves a more extensive airway manipulation, due to the need for bronchoscopy and/or serial dilations during trachea entry and put the patient’s airway in contact with the surgeons from the beginning. These factors can increase aerosolization risks compared with OT, in which there is no contact with the patient’s airway during dissection of the neck and tracheal opening is performed quickly with an incision. Also, aerosolization risks can be mitigated with some specific measures like advancing the tube with the cuff inflated before opening the trachea. Recently, an improved method to perform a PT was proposed. Authors suggests the use of a double lumen endotracheal tube (DLET) for PT in critically ill patients. DLET was equipped with an upper channel that allows passage of a bronchoscope during the percutaneous tracheostomy and with a lower channel exclusively dedicated to patient ventilation. The lower channel is equipped with a distal cuff positioned just above the carina that may allow a safe mechanical ventilation by keeping stable gas-exchange and limiting the spread of aerosol during the procedure.^51,52^

However, authors recommend considering either open tracheostomy or percutaneous tracheostomy at the discretion of the faculty performing the procedure, and according to departmental experience.

#### 7. Operating room or ICU, where is the best place to do it?

All the reviewed guidelines emphasize the importance of a proper surgical setting.^13-29^ Evidence from the SARS-CoV-1 outbreak, highlight the role of bedside tracheostomy in the ICU in negative-pressure rooms.^49,50,53,54^ These measures are directed to avoid unnecessary transport of patients and repeated handling (connection and disconnection) of ventilatory circuits during transfer, which may increase the risk of contamination. Nevertheless, some guideline recommends establishing a COVID-19 Operating room (OR) to perform this procedure. In this case, it should be ideal to do it in a negative pressure OR, with well-demarcated areas and routes for patient transport.^26,29,55^

#### 8. Who should perform the tracheostomy, and which are the required staff during procedure?

Almost all the guidelines recommend minimal staff during the procedure,^13-29^ composed of two surgeons, one anesthetist or ICU doctor, one scrub nurse and a second nurse as a runner. All guidelines recommend a skilled surgical team (COVID-19 Airway team) composed of a senior consultant (ENT or Head & Neck Surgeon) to lead the surgery, assisted by a Skilled assistant (Junior Faculty or consultant), who needs to be familiar with all the surgical steps of tracheostomy in order to minimize the time spent in the contaminated room.^29^

#### 9. What type of dissection is recommended?

Surgical smoke is created when the electrocautery instrument is used to cut, coagulate, or ablate tissue. The tissue is heated to the boiling point of its constituent fluids, causing membranes to rupture, ejecting a bioaerosol.^56^ The use of an efficient, filtered evacuation system is indicated, particularly where there is the possibility of dissemination of viral particles.^57^ Only 6 guidelines mention anything about the use of cold or diathermy dissection. Five guidelines recommend avoiding the use of diathermy.^15,21,23,25,26^ One guideline recommends minimizing the use of diathermy.^24^

#### 10. What are the anesthetic precautions before opening the trachea?

The first step recommended is the suction of oropharyngeal, hypopharyngeal and tracheal secretions with a Yankauer suction tip and an in-line suction system.^21^ Prior to making tracheal window, neuromuscular blockade should be confirmed with the anesthetist in order to avoid swallowing and cough reflexes. Oxygenation must be achieved. Once the anterior wall of the trachea is exposed, the anesthetist or ICU doctor needs: 1) to reduce the oxygen-percentage of the inflated air to 21%, 2)to advance blindly the tube as caudally as possible with cuff inflated, looking for signs of right main bronchial intubation and 3) to hyper-inflate the tube cuff to ensure lower airway isolation and reduce the risk of aerosoling.^58^ Prior to making the tracheal window an additional dose of muscle relaxant can be useful; ventilation needs to be stopped to avoid aerosolization in case of cuff puncture.^21^

#### 11. Post-Tracheostomy care and first change

Post-tracheostomy care should be performed by trained nursing staff. Safe suction with a closed airway circuit and regular checks of cuff-pressure is recommended. No dressing change is recommended unless evidence of local infection, also cannula cuff should not be deflated to avoid any risk of air leakage from the stoma. Recommendation about first change varies among clinical guidelines. Italian guidelines suggest that the first change should be planned 7-10 days later using the same standards (PPE utilization and airflow interruption), and subsequent cannula change can be delayed 30 days after.^26^ Spanish Guidelines suggest waiting 14-21 days.^23^ British guidelines from the BLA suggest waiting almost 4 weeks,^21^ by contrast ENT-UK guideline suggests making the first change in the first 7-10 days.^22^ South-African and the AAO-HNS guidelines suggests delaying routine post-operative tracheotomy tube changes until COVID-19 testing is negative.^15,27,28^ Other North American guideline suggest avoiding tube changes unless clinically indicated.^17^

#### 12. Filters and protective measures in tracheostomy patient

All guidelines suggest using Humidity-Moisture filters (HME) after tracheostomy.^13-30^ Commercial filters are described and recommended for laryngectomy patients and for patients after elective tracheostomy given many carriers are asymptomatic. Both products have an effective electrostatic filter that can protect against >99% of viruses and bacteria according to the composition.

### Study limitations

The present study has some limitations. The absence of prospective data about indications, timing and relevance of tracheostomy in COVID-19 patients is limiting the level of evidence used to provide robust guidelines. We have probably not included all the existing guidelines since many of them are not available online.

## Conclusions

Tracheostomy represents a high-risk AGP due to constant exposure to droplets and aerosol leakage infected with of SARS-CoV-2 during the surgical procedure. Full PPE is mandatory and the creation of a COVID-19 airway team is essential. Otolaryngologist - Head & Neck Surgeons performing tracheostomies should be aware that they are at the highest risk category together with emergency doctors, anesthesiologist and ICU nurses and doctors. Collecting data about indications, clinical outcomes and safety for health providers of tracheotomies performed in the setting of SARS-CoV-2 outbreak will help building more robust evidence-based recommendations.

### YO-IFOS tracheostomy recommendations

- All cases should ideally be discussed in a multidisciplinary team, to evaluate the risks vs benefits of the procedure for the patient and the entire health care team.
- Intensive hand hygiene with hydroalcoholic solution. Full PPE due to the high risk of infection, including N95 or N99 (USA), FFP2 or FFP3 (Europe) mask, the use of double gloves, goggles or eye protection, PARP, face shield and an apron or gown is recommended in al suspected or positive patients.
- Elderly, hypertension, diabetes, chronic obstructive pulmonary disease, cardiovascular disease, and cerebrovascular disease are significant risk factors for COVID-19 patients.
- Not enough evidence is available to suggest the ideal time for tracheostomy. However, available evidence suggests considering tracheostomy after 14 days of invasive mechanical ventilation.
- Consider either open tracheostomy or percutaneous tracheostomy according to the faculty performing the procedure and the institution resources.
- Bedside tracheostomy in the ICU or in Operative Room with negative pressure.
- Establish a specific COVID-19 Airway team.
- Minimizing or avoiding the use of diathermy is recommended.
- Follow a Minimal-Staff policy on each tracheostomy.
- HME are mandatory after tracheostomy.
- Delay first tracheal change.

## Data Availability

Not apply

## Notes

### Competing Interest Statement

The authors have declared no competing interest.

### Funding Statement

No funding was required for this research

